# Artificial Intelligence in Mammography Screening in Norway (AIMS Norway): Protocol for a randomized controlled trial

**DOI:** 10.64898/2026.03.13.26348320

**Authors:** Åsne Sørlien Holen, Marthe Larsen, Solveig Hofvind

**Affiliations:** Department of Breast Cancer Screening, Cancer Registry of Norway, Norwegian Institute of Public Health, P.O. Box 5313 Majorstuen, 0304 Oslo, Norway; Department of Health and Care Sciences, Faculty of Health Sciences, UiT The Arctic University of Norway, P.O. Box 6050 Stakkevollan, 9037 Tromsø, Norway

## Abstract

**Background and Objective:** Increasing screening volumes, combined with global shortage of radiologists and a high proportion of normal mammograms, challenge the efficiency and sustainability of breast cancer screening. Artificial intelligence (AI) has the potential to improve resource allocation, workflow efficiency and diagnostic performance by supporting and partially replacing radiologists in the interpretation process. This randomized, controlled, parallel-group, non-inferiority, single-blinded trial evaluates whether an AI-supported reading strategy, involving one or two radiologists depending on AI risk stratification, is non-inferior to standard independent double reading. The primary outcome is the number of screen-detected breast cancer cases in each group.

**Methods:** Women invited to BreastScreen Norway in the Western, Central, and Northern Norway Regional Health Authorities are eligible for inclusion. Following written informed consent, participants are randomized 1:1 to the control group (standard independent double reading by two radiologists) or the intervention group. In the intervention group, mammograms are analyzed using Transpara. Examinations with AI scores of 1–7 are interpreted by a single radiologist, whereas examinations with scores of 8–10 undergo independent double reading. Radiologists are blinded to AI scores and AI image markings during the initial interpretation; this information is disclosed during consensus meetings. Non-inferiority will be assessed by estimating confidence interval for the difference in screen-detected cancer rates between groups. Non-inferiority will be concluded if the upper bound of the confidence interval does not exceed the predefined non-inferiority margin.

**Conclusions:** The trial addresses a critical challenge in breast cancer screening: maintaining diagnostic performance while improving efficiency in the context of workforce constraints and a high prevalence of normal examinations. By evaluating a risk-stratified AI-supported reading strategy within a population-based screening program, the study will provide important evidence on whether AI can be safely integrated to optimize workload distribution while preserving cancer detection rates.

**Trial registration:** The ClinicalTrials.gov registry (NCT06032390)

## Introduction

### Background

Breast cancer is a major global health concern, with over half a million women dying from the disease annually (1). In response, many countries, including Norway, have implemented mammographic screening programs to enable early detection. These programs, along with improved treatment, have resulted in a substantial reduction in breast cancer mortality among screening participants (2).

In Norway, the national screening program, BreastScreen Norway, invites all women aged 50-69 to two-view biennial mammography (3). Attendance is approximately 77% of the target population of 680 000 women in each screening round. Mammograms are interpreted using independent double reading with consensus, as recommended in European guidelines (4). Radiologists assign each breast a score from 1 to 5: 1, negative; 2, probably benign; 3, intermediate suspicion; 4, probably malignant; 5, high suspicion of malignancy (3). Cases scored 2 or higher by either radiologist are discussed in a consensus meeting which includes at least two radiologists to determine whether recall is warranted. Women recalled undergo further diagnostic procedures. The consensus rate is about 9%, the recall rate about 3% and approximately 0.6% of all screenings results in a breast cancer diagnosis, meaning that over 99% of examinations are ultimately classified as negative.

Global shortage of radiologists (5–8), and the high volume of predominantly negative examinations pose challenges for workflow efficacy and resource allocation. Moreover, studies have shown that nearly one quarter of the screen-detected cancers are initially interpreted as negative by one of the two radiologists (9–11). Retrospective analyses further indicate that 20-25% of both screen-detected and interval cancers (breast cancer diagnosed between two screening episodes on the basis of symptoms) are retrospectively visible on prior mammograms, highlighting the inherent limitations and subjectivity of mammographic interpretation (12–14).

Artificial intelligence (AI) has emerged as a promising approach to address these challenges. Deep learning, in particular, has been applied across healthcare, including radiology, to improve efficiency and augment clinical decision making (13). Retrospective evaluations of AI in mammography have demonstrated strong performance in distinguishing cancer-positive from cancer-negative examinations (15–23). These systems generate malignancy risk scores, reflecting the estimated probability of cancer with higher scores indicating greater suspicion of malignancy.

### Retrospective evidence

Several studies aimed at evaluating AI’s ability to detect cancers retrospectively; compare performance with independent double reading; assess performance across breast density categories; explore AI-based triaging; analyze imaging characteristics of true- and false-positive findings; and assess whether AI markings correspond to actual tumor locations, have been performed on retrospective data from BreastScreen (9, 24–27).

High AI scores were strongly associated with both screen-detected and interval cancers. Using a 1-10 scale, where 10 indicated high risk of breast cancer, 87-90% of the screen-detected and about 45% of the interval cancers were assigned with an AI score of 10 (9, 25). Sensitivity at score 10 was ~80%, increasing to >90% for scores 8–10 (25). AI matched or exceeded individual radiologists’ performance flagging many cancers missed during independent reading (24). Triaging low-risk examinations with AI could reduce radiologist workload, with up to 70% of exams classified as low risk without substantially affecting cancer detection. AI lesion localization was highly accurate for screen-detected cancers and moderately accurate for interval cancers with high scores (26), though many interval cancers had subtle or no visible signs, limiting earlier detection.

High scores indicate suspicion, but do not establish diagnosis – breast radiologists’ evaluation and histopathological confirmation remain essential. Prospective studies are therefore required to evaluate AI’s real-world performance, including optimal integration strategies, thresholds for replacing a radiologist, and impacts on consensus decisions, recall rates, cancer detection, and workload.

### Objectives

Based on the mentioned knowledge gaps, we designed a randomized controlled trial (RCT) to evaluate AI assisted interpretation of screening mammograms within BreastScreen Norway. This randomized, controlled, parallel-group, non-inferiority, single-blinded trial aims to compare the number of breast cancers detected at screening among participants whose mammograms were triaged to low or intermediate/high risk using AI and interpreted by one or two radiologists, respectively, versus those whose mammograms were interpreted using standard independent double reading.

The primary objective is to determine whether AI-assisted interpretation is non-inferior to the current standard, independent double reading. Specifically, the trial seeks to answer: Can independent double reading be safely replaced by an AI-assisted strategy that triages examinations according to AI-generated cancer risk scores, without compromising the breast cancer detection rate?

## Methods

The protocol was developed and reported in accordance with the SPIRIT (Standard Protocol Items: Recommendations for Interventional Trials) guidelines to ensure completeness and transparency.

### The AI model

The trial will use Transpara® (ScreenPoint Medical AB, the Netherlands,) a CE marked Class IIb device, for automated mammography interpretation (28). Transpara® employs convolutional neural networks and is trained on mammograms from multiple screening programs and vendors (17, 29). It generates a continuous malignancy probability (raw score) and an overall exam-level AI score ranging from 1 to 10, calibrated to allocate approximately 10% of exams to each score. Scores of 1 indicate low probability of malignancy, whereas scores of 10 indicates high probability. Scores 1-7 are low risk, 8-9 intermediate risk, and 10 high risk. The Sectra Amplifier Platform facilitate image analyses (30).

### Recruitment and consent

All women invited to BreastScreen Norway in the Western, Central, and Northern Regional Health Authorities are eligible for inclusion. Trial information is included with the screening invitation letter; an English version is available online and at the screening sites.

During the standard pre-screening interview, radiographers invite participants and obtain written informed consent. Participation is voluntary and can be withdrawn at any time without affecting current or future screening attendance. Screening occurs at 12 stationary and four mobile screening units with screen-reading, consensus and further assessment being performed at eight designated breast centers. Only one has prior clinical AI experience. Approximately 100,000 women are screened annually across these regions, with an average screen-detected breast cancer rate of 6.1 per 1,000 examinations (31).

### Randomization

Consenting participants are randomized 1:1 to study or control group immediately after the pre-screening interview via a computer-generated algorithm integrated into the screening IT system, based on the woman’s personal identification number (Figure 1). Allocation is concealed from participants and radiographers. All participants undergo standard CC and MLO digital mammography. Only the interpretation procedure differs between the groups, which constitutes the intervention being studied.

**Figure 1:**
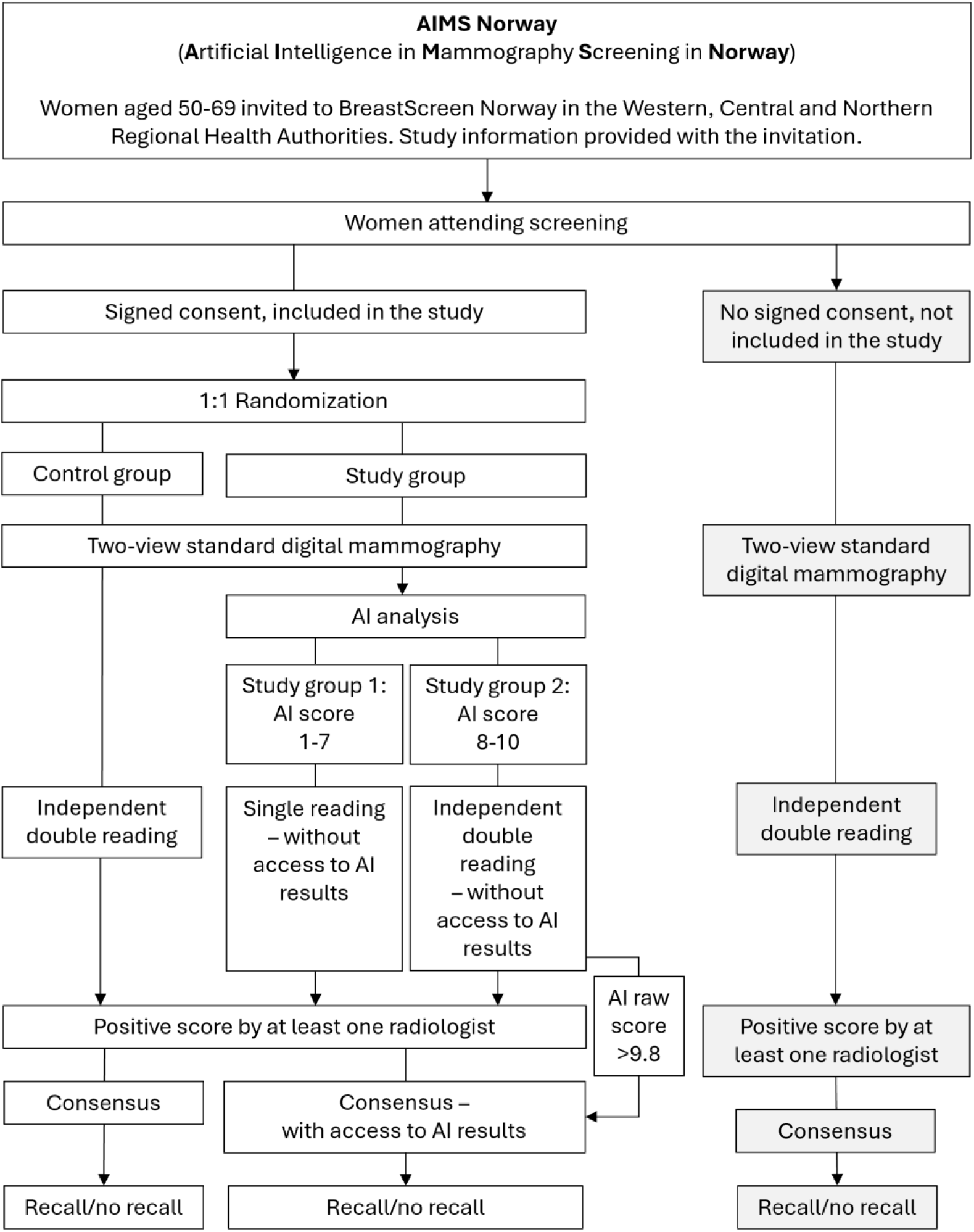
Trial workflow diagram

### Control group

Control group participants receive standard independent double reading by two radiologists, with access to prior images and consensus meetings for flagged cases (3, 10). Follow-up and recall examinations are per standard procedures. AI analysis will be performed retrospectively after recruitment has been concluded, for research purposes only.

### Study group

Mammograms in the study group will be analyzed using Transpara® prior to interpretation. Based on the AI-generated malignancy risk score:

- AI scores 1–7: Single reading by radiologist (Study group 1)
- AI scores 8–10: Independent double reading (Study group 2)

Radiologists are blinded to AI scores and CAD marks during initial reading and are unaware of whether the exam is Study group 1 or 2. AI-processed images include a non-removable watermark, and the radiologist knows whether they are reading an examination in the study or control group. All cases flagged by radiologists or with an AI raw score of >9.8 undergo consensus review, where AI scores and CAD marks are disclosed. Prior screening images are available per standard practice.

### Power and sample size calculations

The primary endpoint is screen-detected cancer (DCIS or invasive). Assuming a control group rate of 6.1 cancers per 1,000 exams, the non-inferiority margin is set to 0.0012, using 4.9 cancers per 1,000 as the lowest acceptable threshold. With 80% power and α = 0.025, 132,184 exams (66,092 per arm) are required to prove non-inferior cancer detection between the groups. Assuming 80% participation, 165,230 women must be invited to participate in the study, which is achievable during one screening round (2 years). Table 1 presents sample size calculations across participation rates and power levels.

**Table 1:** Sample size (n) calculation for different participation rates in the trial among those attending the screening program – acceptance rate, by different beta (β) values, using a non-inferiority margin of 0.0012 and alfa (α) = 0.025.

| Beta | Number of women required in the trial to demonstrate non-inferiority for screen-detected cancers | Estimated number of women to be invited for trial participation based on varying acceptance rates (70–90%) |  |  |  |  |
| --- | --- | --- | --- | --- | --- | --- |
|  |  | 90% | 85% | 80% | 75% | 70% |
| $\beta=0.8$ | n=132,184 | n=146,872 | n=155,511 | **n=165,230** | n=176,246 | n=188,835 |
| $\beta=0.9$ | n=176,958 | n=196,620 | n=208,186 | n=221,198 | n=235,944 | n=252,798 |

### Study outcomes

#### Primary outcome

Number of screen-detected breast cancers.

#### Secondary outcomes

Consensus rate, recall rate, interval cancer rate, tumor histopathology, mammographic features, and time spent on reading and consensus meetings. Interval cancer is defined as breast cancer diagnosed within 24 months after a negative screen or 6–24 months following a false-positive screen (4).

### Data collection and analysis

Screening and interpretation data are stored in dedicated databases at BreastScreen Norway and the Cancer Registry of Norway (NIPH), undergo quality control, and are merged with AI output data. Data is pseudonymized prior to analysis. Non-inferiority will be tested with confidence interval of the difference in screen-detected cancer between the control- and study group. The null hypothesis is rejected if the upper limit is below the non-inferiority margin. Descriptive statistics summarize participant characteristics. Analyses will be conducted in Stata.

### Monitoring

A Data Monitoring and Safety Board (DSMB) oversees trial integrity, participant safety, and quality, reviewing interim results every six months. The trial research group at the NIPH ensures data quality and monitors performance metrics. Persistent deviations are reported to the DSMB, which may recommend trial termination. An expert group of radiologists and lead contacts meets biweekly, and research radiographers convene monthly to review operational aspects.

### Ethics and dissemination

AIMS Norway has received ethical approval from the Regional Committees for Medical and Health Research Ethics (REC; reference no. 366405). Furthermore, the Cancer Registry of Norway, at the Norwegian Institute of Public Health has conducted a Data Protection Impact Assessment (DPIA) for the study.

All participants will provide written informed consent before enrolment. This consent constitutes the legal basis for the processing of personal data in accordance with the General Data Protection Regulation (GDPR), pursuant to Article 6(1)(a) for personal data and Article 9(2)(a) for special categories of personal data. The consent includes permission to collect and process study-specific data as well as relevant screening information, including data from the index examination and any subsequent follow-up procedures and outcomes. Participants may withdraw their consent at any time without consequences. Upon withdrawal, all study-specific data not yet anonymized will be deleted from the study database. However, data that has already been anonymized and incorporated into analyses or publications cannot be removed. Participants also have the right to access information about their personal data, including how the data are processed and who has accessed the data.

All study data will be collected and stored in secure, dedicated databases in accordance with established procedures at BreastScreen Norway and the Cancer Registry of Norway, NIPH. Prior to statistical analyses, all data will be pseudonymized to protect participant confidentiality.

De-identified trial data may be shared upon reasonable request, in accordance with the data processing principles set out in Article 5 of the GDPR, provided that the applicant demonstrates an appropriate legal basis under Articles 6(1)(e) and 9(2)(j), as well as supplementary authorization under applicable Union or Member State law and approval from REC. Any transfer of data to third country or international organizations will comply with the requirements set out in Chapter V of the GDPR.

The full study protocol (Version 9, dated October 6, 2025) and the statistical analysis plan (Version 5, dated October 6, 2025) may be obtained by contacting the project leader at the Norwegian Institute of Public Health.

The study findings will be disseminated through publication in international, open-access, peer-reviewed journals within the fields of radiology, artificial intelligence, and epidemiology. In addition, lay summaries will be made publicly available via the Norwegian Institute of Public Health’s website to ensure accessibility for participants, stakeholders, and the public.

## Discussion

This randomized, controlled, non-inferiority trial is designed to evaluate whether an AI-supported reading strategy in population-based mammography screening can maintain breast cancer detection while reducing reliance on independent double reading by radiologists. The study addresses a key operational challenge in contemporary screening practice. Increasing screening volumes, combined with a high proportion of normal examinations and persistent shortages of specialized breast radiologists, place substantial pressure on screening programs and may threaten their long-term sustainability.

Artificial intelligence has the potential to improve workflow efficiency by prioritizing radiologist attention toward examinations with a higher likelihood of malignancy. In this trial, AI is used to stratify mammograms according to risk: examinations classified as low risk are interpreted by a single radiologist, whereas examinations classified as intermediate or high risk continue to undergo independent double reading. This approach reflects the distribution of findings in screening populations, where most examinations are normal, and where efficiency gains are therefore most likely to be realized. To minimize the risk of automation bias, AI scores and image markings are not available during initial image interpretation and are disclosed only during the consensus process. This design feature enhances the internal validity of the study and supports an unbiased evaluation of the non-inferiority hypothesis.

A non-inferiority design was selected to determine whether reducing the level of independent human reading does not result in a clinically unacceptable reduction in cancer detection. The screen-detected breast cancer rate is a key performance indicator in mammography screening and represents the most directly relevant outcome for evaluating the safety of reduced radiologist involvement. However, cancer detection alone does not capture the full impact of a modified reading strategy. Secondary outcomes, including interval cancer rates, recall rates, false-positive findings, and tumor characteristics, will be essential to provide a comprehensive assessment of the balance between screening effectiveness and efficiency, and to identify any unintended consequences.

Overall, this trial will generate robust evidence on whether a risk-stratified, AI-supported reading strategy can safely reduce dependence on independent double reading in mammography screening. The findings are expected to inform future screening policies and may contribute to the development of more efficient and sustainable screening models that appropriately integrate AI while preserving the benefits of human expertise.

## Data Availability

All data produced in the present work are contained in the manuscript.

## Ethics approval

The Regional Committee for Medical and Health Research Ethics in South-Eastern Norway gave ethical approval for this work (#366405).

